# Phenomenological Modelling of COVID-19 epidemics in Sri Lanka, Italy and Hebei Province of China

**DOI:** 10.1101/2020.05.04.20091132

**Authors:** A.M.C.H. Attanayake, S.S.N. Perera, S. Jayasinghe

**Affiliations:** Department of Statistics & Computer Science, Faculty of Science, University of Kelaniya.; Research & Development Centre for Mathematical Modelling, Department of Mathematics, Faculty of Science, University of Colombo.; Department of Clinical Medicine, Faculty of Medicine, University of Colombo

**Keywords:** Novel Coronavirus (COVID-19), Phenomenological Models, Epidemiological Curve, Prediction

## Abstract

The COVID-19 pandemic has resulted in increasing number of infections and deaths on a daily basis. There is no specific treatment or vaccine identified and the focus has been preventive measures based on statistical and mathematical models. These have relied on analyzing the behavior of populations and characteristics of the infection and applying modelling techniques. The analysis of epidemiological curve fitting on number of daily infections across affected countries could give useful insights on the characteristics of the epidemic. A variety of phenomenological models are available to capture dynamics of disease spread and growth. Data for this study used the number of daily new infections and cumulative number of infections in COVID-19 in three selected countries, Sri Lanka, Italy and Hebei province of China, from the first day of appearance of cases to 20^th^ April 2020. In this study Gompertz, Logistic and Exponential growth curves were fitted on cumulative number of infections across countries. Akaike’s information criteria (AIC) was used in determining the best fitting curve for each country. Results revealed that the most appropriate growth curves for Sri Lanka, Italy and China-Hebei are Exponential, Gompertz and Logistic curves respectively. The overall growth rate and final epidemic size evaluated from best models for the three countries and short-term forecasts were also generated. Log incidences over time in each country were regressed before and after the identified peak time of the respective outbreaks of countries. Hence, doubling time/halving time together with daily growth rates and predictions were estimated. Findings altogether demonstrate that outbreak seems extinct in Hebei-China whereas further transmissions are possible in Sri Lanka. In Italy, current outbreak transmits in a decreasing rate.

## 1. Introduction

COVID-19 or novel coronavirus is a pandemic where initiated in Wuhan, China in December 2019. At present the pandemic was recorded total number of 2,471,136 infections and 169,006 deaths in the world [1]. Daily infections and deaths are increasing at an alarming rate. According to the situation report 93 of WHO, total confirmed cases were 84,287 and deaths were 4,642 in China as at 22 April 2020. These figures for Italy and Sri Lanka were 183,957 and 24,648 and 310 and 7 respectively. On 19^th^ March 2020 Italy was the country where the highest number of deaths were recorded among the other countries in the world. Since then Italy has attracted the attention of the public on its active engagement with the coronavirus. In Sri Lanka, the first infection was identified on 11^th^ March 2020. Afterwards as of other countries several control measures such as isolation of cases, quarantine of suspects, lockdown of the country or permitting island wide curfew, limitation of public gatherings, annunciation of social distancing, increase awareness of the disease through media and encouraging people to stay in their homes were implemented. Clearly, China was at the phase of ending epidemic when Italy entered to the phase of a growing epidemic in February 2020 and now is in its middle phase. Sri Lanka had the beginnings of the epidemic in March 2020 and is still in the early phase. Since these three countries are in the three phases of the epidemic, we decided to analyze them further and draw some conclusions.

To mitigate the epidemic control measures should be coupled with mathematical and statistical modelling to fine tune strategies by anticipating the dynamics of disease spread. Applications of phenomenological models in describing a growth of natural phenomenon can be found in the literature [2, 3, 4, 5]. These models are able to capture empirical patterns of an epidemic and are useful in predicting dynamics of the disease. Prediction of models will be beneficial for health care professional to allocate necessary resources and implement preventive measures at the appropriate time.

The analysis of epidemiological curve fitting on number of daily infections provides a useful insight on determination of the variety of statistical features of an outbreak [6,7]. To capture the epidemic pattern one can fit a regression model on log incidences over time. After the peak of the distribution has identified pre and post peak models of the distribution lead to identify the point of zero incidences in an outbreak. Further, the estimation of doubling time or halving time together with daily growth rates and predictions were supported by the curve fitting.

In this study Gompertz, Logistic and Exponential growth curves fitted on cumulative number of infections over the selected three countries; Sri Lanka, Italy and Hebei province of China and the best fitted growth curve was identified for each country. The overall growth rate and final epidemic size evaluated from best models for the three countries and short-term forecasts were also generated. Further, useful characteristics of epidemiological curves were extracted and predictions were generated using pre and post peak models.

## 2. Materials and Methods

### 2.1 Data

Daily accumulated COVID-19 cases were downloaded [8] for the three countries from the first day of appearance of cases to 20^th^ April 2020. Data recording began in Hebei province of China on 22^nd^ of January 2020 and in Italy on 31^st^ of January 2020. That is for Sri Lanka was on 10^th^ of March 2020. Daily new infections were calculated from the downloaded data.

### 2.2 Methods and Definitions

#### 2.2.1 Exponential Growth Model

Exponential growth models are suitable in fitting early phase of an epidemic because it is not realistic to represent a natural growth by an exponential model which may never ends. An exponential growth explained as:

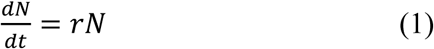

with the solution

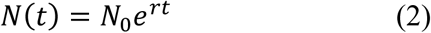

where *N*_0_ denotes the size of the population at time zero, *N* denotes the size of the population, *t* denotes time and *r* is the per capita rate of increase.

#### 2.2.2 Logistic Growth Model

Logistic growth models have approximately exponential nature at the first phase and continue the growth at reduced rate of growth where finally reach to its maximum. The mathematical model that explains the logistic growth expressed as:

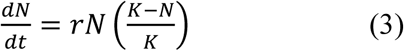

where *N* denotes the size of the population, *t* denotes time, *K* is the carrying capacity and *r* is the per capita growth rate.

The solution of (3) is given:

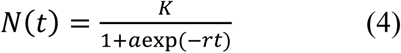

where 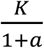 is the initial value.

#### 2.2.3 Gompertz Growth Model

Gompertz model is a sigmoid function which describes slowest rate of growth at the beginning and at the end. The model can be written as:

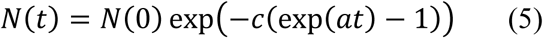

where *a* is the rate of growth, *N*(0) is the initial number of organism, *c* denotes the displacement across the X-axis and *b* and *c* are positive numbers.

#### 2.2.4 Akaike Information Criteria (AIC)

AIC is calculated by using the formula:

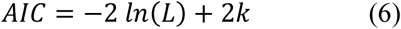

where *k* is the number of parameters in the model and *L* be the maximum value of the likelihood function for the model. A model with the minimum AIC value will be the best model.

#### 2.2.5 Epidemic Curve

Epidemiological curve or an epidemic curve is a curve use in epidemiology to extract meaningful characteristics of an epidemic. Specially, it shows the transmission of the disease over time. Further, disease outliers, magnitude of the epidemic, its peak time, applicable trends and incubation period identified by the curve.

#### 2.2.6 Growth Rate

Growth rate denotes the change of a particular variable over a specified time duration. This statistic is useful to describe the performance of a variable as well as to predict the performance. The growth rate calculated as follows:

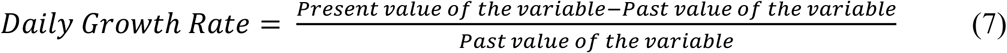

The growth rate over *n* time duration is given by:

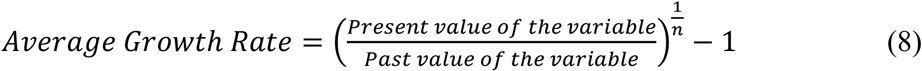

## 3. Results and Discussion

R software is used in the analysis [9]. The number of daily incidences of COVID-19 in Hebei province of China is shown in Figure 1 and accumulated incidences in Figure 2. In between January and March of 2020 the number of daily incidences varies with the maximum value of 23 cases. Figures illustrate that after February of 2020 number of reported incidences were lower in Hebei and epidemic reaches its saturation.

**Figure 1:**
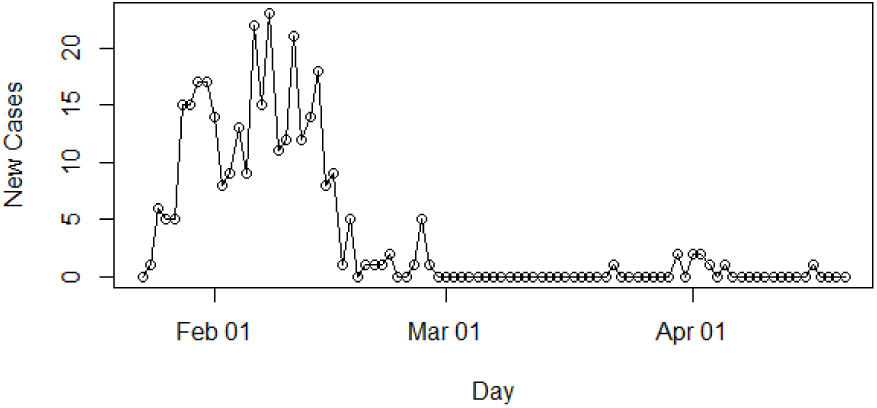
Daily Incidences of Coronavirus in Hebei

**Figure 2:**
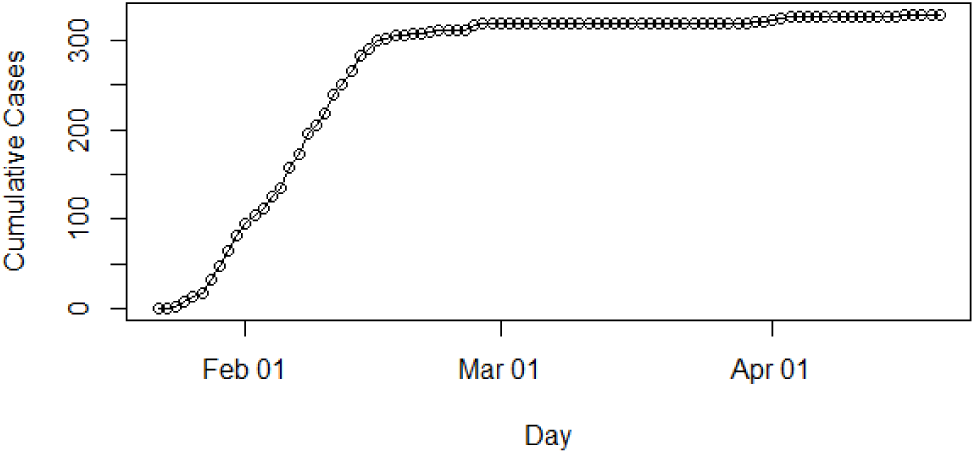
Cumulative Incidences of Coronavirus in Hebei

The daily number of new infections in Italy is depicted in Figure 3 and its cumulative distribution in Figure 4. Slow transmission is apparent at the beginning stage of the epidemic. The daily incidences in Italy ranges in between 0 and 6,557 cases which has mean of 2,265 cases with standard error of mean of 245. According to Figure 4, epidemic is downgrading but still not reaches its saturation. Therefore, more cases can be expected under reduced rate.

**Figure 3:**
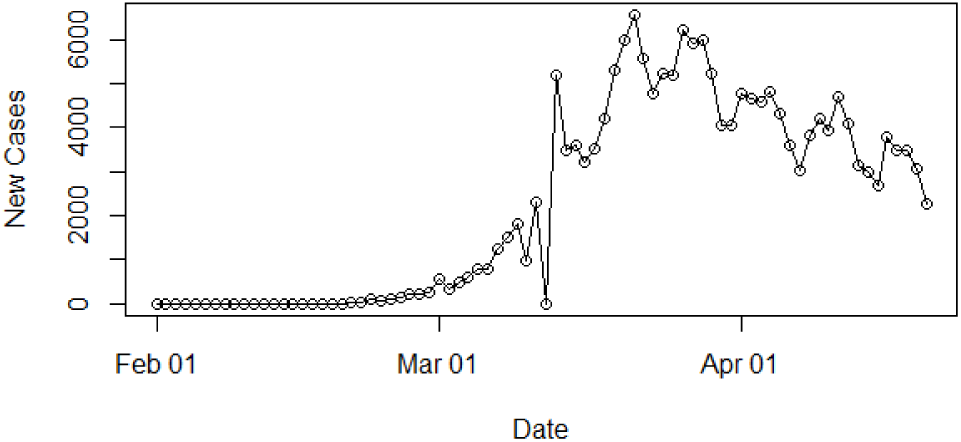
Daily Incidences of Coronavirus in Italy

**Figure 4:**
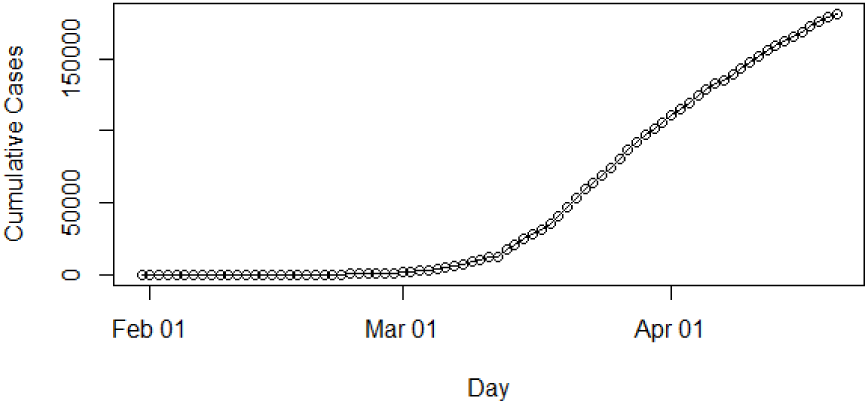
Cumulative Incidences of Coronavirus in Italy

In Sri Lanka, the number of daily recordings of COVID-19 cases represented in Figure 5. The highest number of recording was reported in the 20^th^ of April 2020 which was 33 cases. The date is the final point of the data analysis of this study. Some zig-zag pattern is apparent in the distribution of Figure 5. The cumulative number of infections in Sri Lanka is shown in Figure 6. It is apparent that epidemic is still progressing in Sri Lanka as of date 20^th^ April 2020.

**Figure 5:**
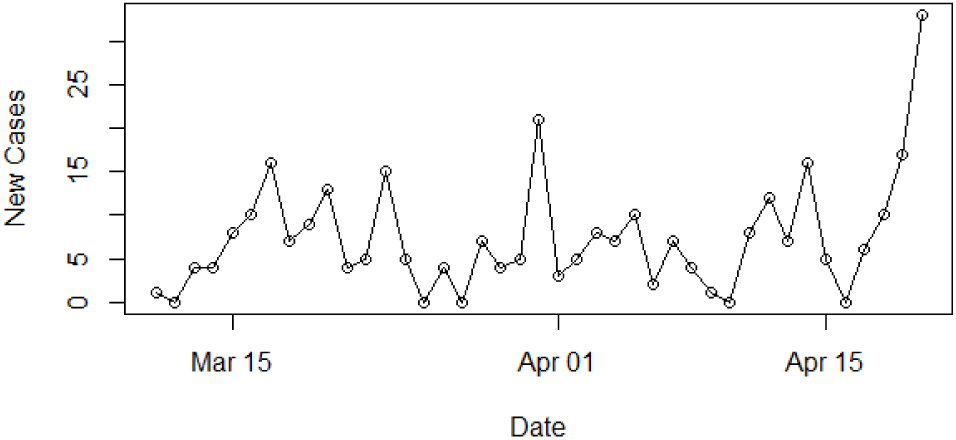
Daily Incidences of Coronavirus in Sri Lanka

**Figure 6:**
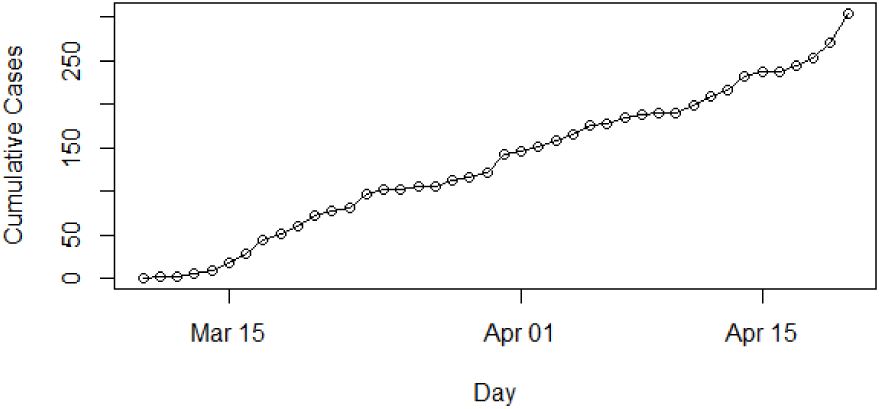
Cumulative Incidences of Coronavirus in Sri Lanka

The epidemiological curve for Hebei, China in Figure 7 shows the number of cases increased till mid of February 2020 and then progressively decreased. This was an indication that the peak time of the outbreak in China had been reached on 8^th^ February 2020.

**Figure 7:**
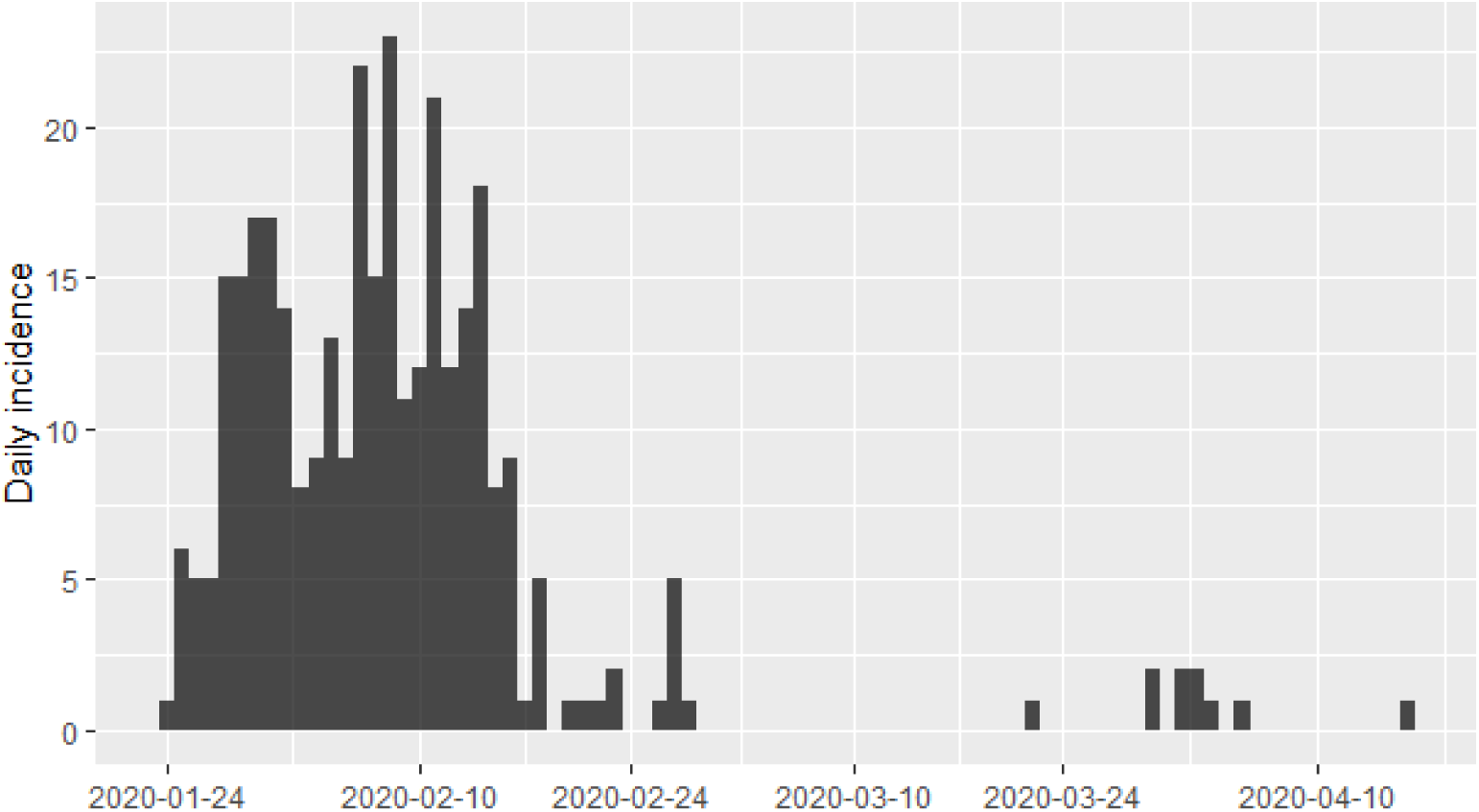
Epidemic Curve of COVID-19 Infections in Hebei Province, China

We then divided the entire data period into two halves as before and after the peak point and regressed log incidences over time for each split. The fitted models are exponential curves as depicted in Figure 8 and 9. Prior to 8^th^ February 2020 the number of incidences were increased at the rate of doubling in every 6 days (95% CI, [3.86, 17.49]) and the daily growth rate was estimated as 0.11 with 95% confidence interval (0.04, 0.18). According to the fitted log-linear model, after the 8^th^ February 2020 the reported incidences were downgraded at the rate of halving in every 20 days (95% CI, [13.13, 4.65]) with a daily decreasing rate of −0.034 (95% CI, [−0.053 −0.016]). Predictions of post peak log-linear model indicated the epidemic almost reached to zero point of infections.

**Figure 8:**
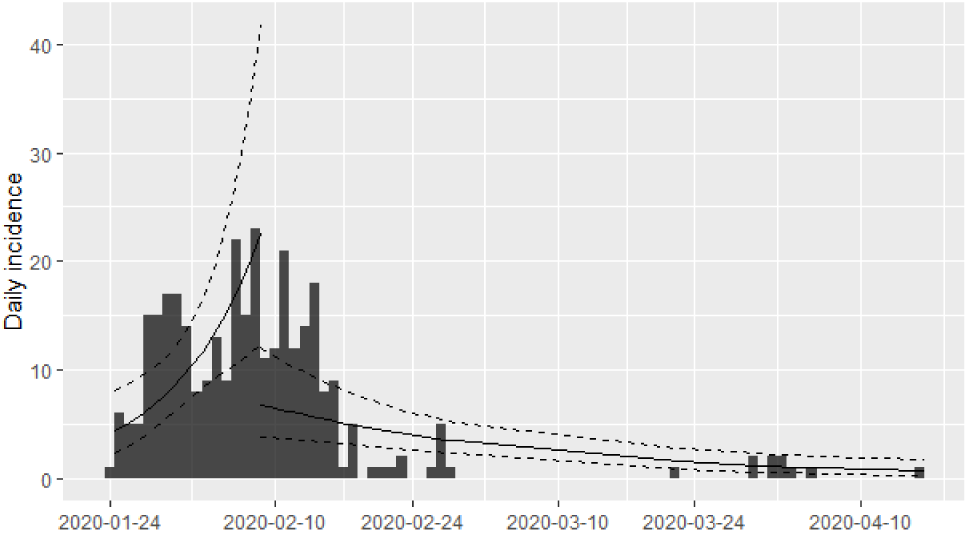
Prediction Models for COVID-19 Infections in Hebei Province, China

**Figure 9:**
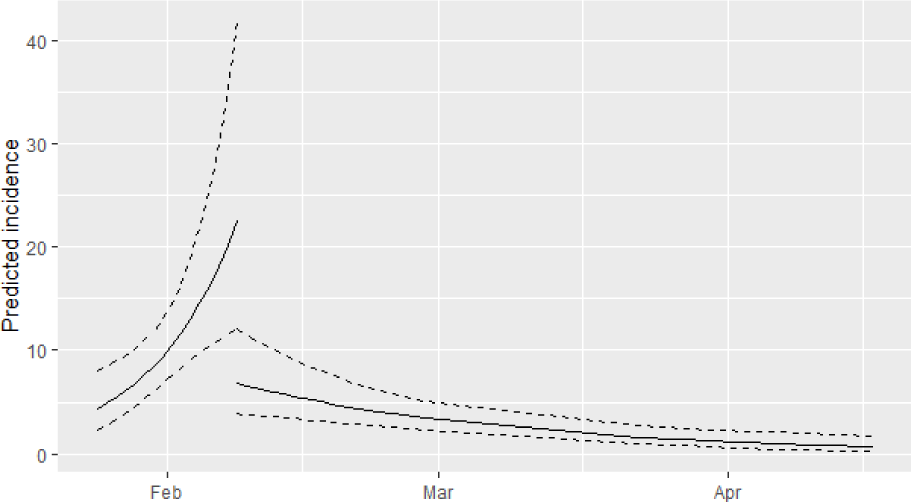
Prediction Models with Intervals

The epidemiological curve for Italy is shown in Figure 10. It can be concluded that the peak time of the outbreak in Italy had reached as apparent growth and shrinkage patterns of the epidemiological curve. The estimated peak time was on 21^st^ March 2020 and the pre and post peak data were separated and each split was regressed by log incidences over time. The fitted exponential curves shown in Figure 11 and 12. From the first date of appearance of cases to 21^st^ March 2020 the number of COVID-19 infections were increased at the rate of doubling in every 4 days (95% CI, [3.27, 3.92]). The daily growth rate was estimated as 0.19 (95% CI, [0.18, 0.21]). After the 21^st^ March 2020, the reported COVID-19 infections rate decreased more slowly, halving every 29 days (95% CI, [23.18, 37.78]). The daily decreasing rate was −0.024 (95% CI, [−0.03, −0.018]). Predictions of post peak log-linear model indicated that the COVID-19 epidemic in Italy is progressing with a decreasing rate.

**Figure 10:**
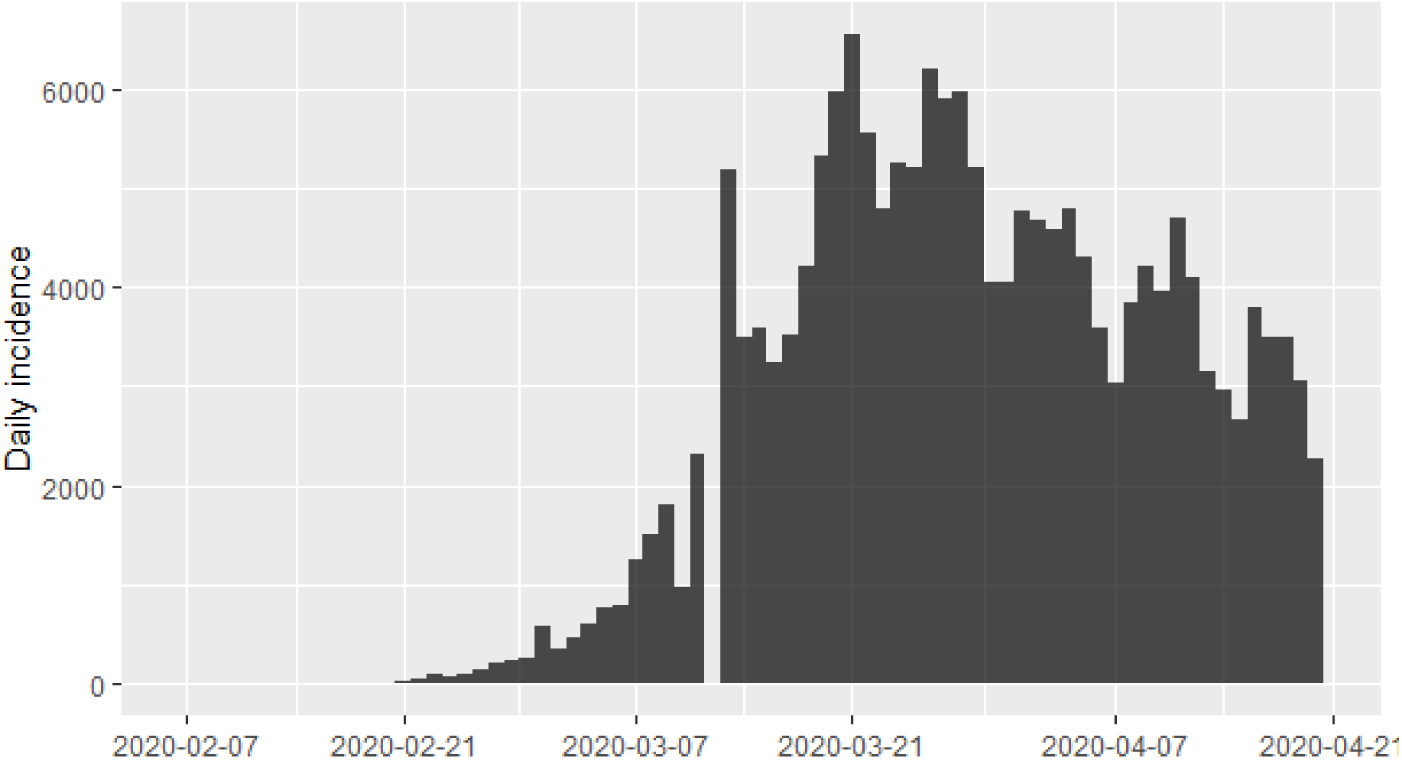
Epidemic Curve of COVID-19 Infections in Italy

**Figure 11:**
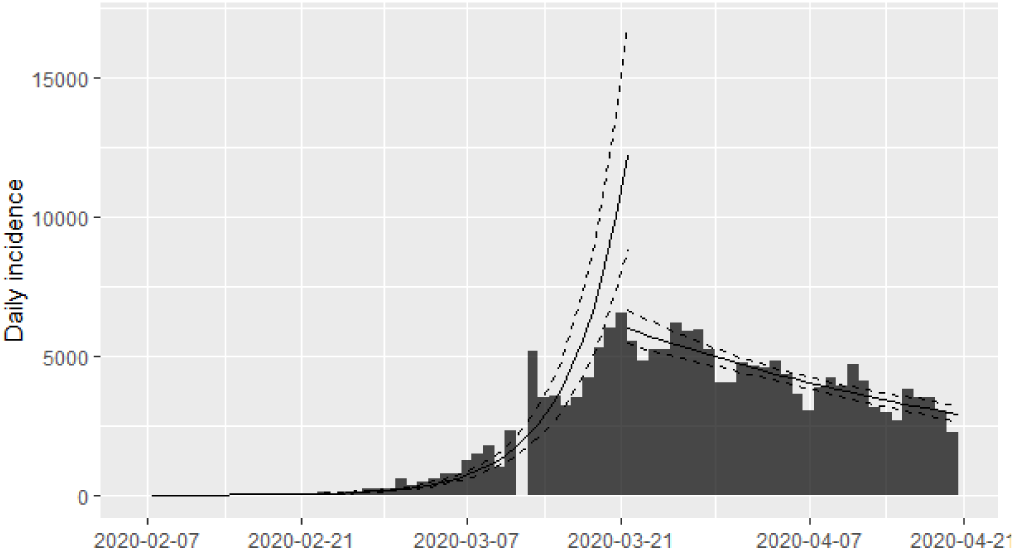
Prediction Models for COVID-19 Infections in Italy

**Figure 12:**
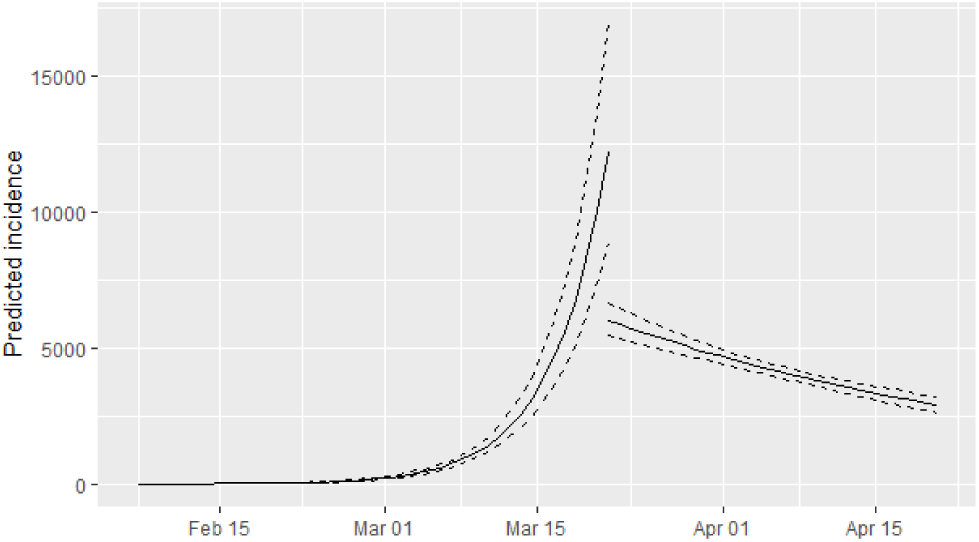
Prediction Models with Intervals

The Figure 13 shows the epidemiological curve for Sri Lanka. Clearly, Sri Lanka is at the initial stage of the coronavirus outbreak. The pattern of the epidemiological curve is undulating and it is a challenge to determine the peak of the distribution. The estimated peak time was on 20^th^ April 2020 which was the last date of the data gathering of this study. This suggested that the outbreak is rapidly increasing. Therefore, only one log-linear model was fitted and shown in Figure 14 and 15. The number of COVID-19 infections is increasing at the rate of doubling in every 45 days. The daily growth rate was estimated as 0.015 with 95% confidence interval (−0.006, 0.037). Predictions of the fitted log-linear model indicated that the epidemic is progressing in Sri Lanka with an increasing rate.

**Figure 13:**
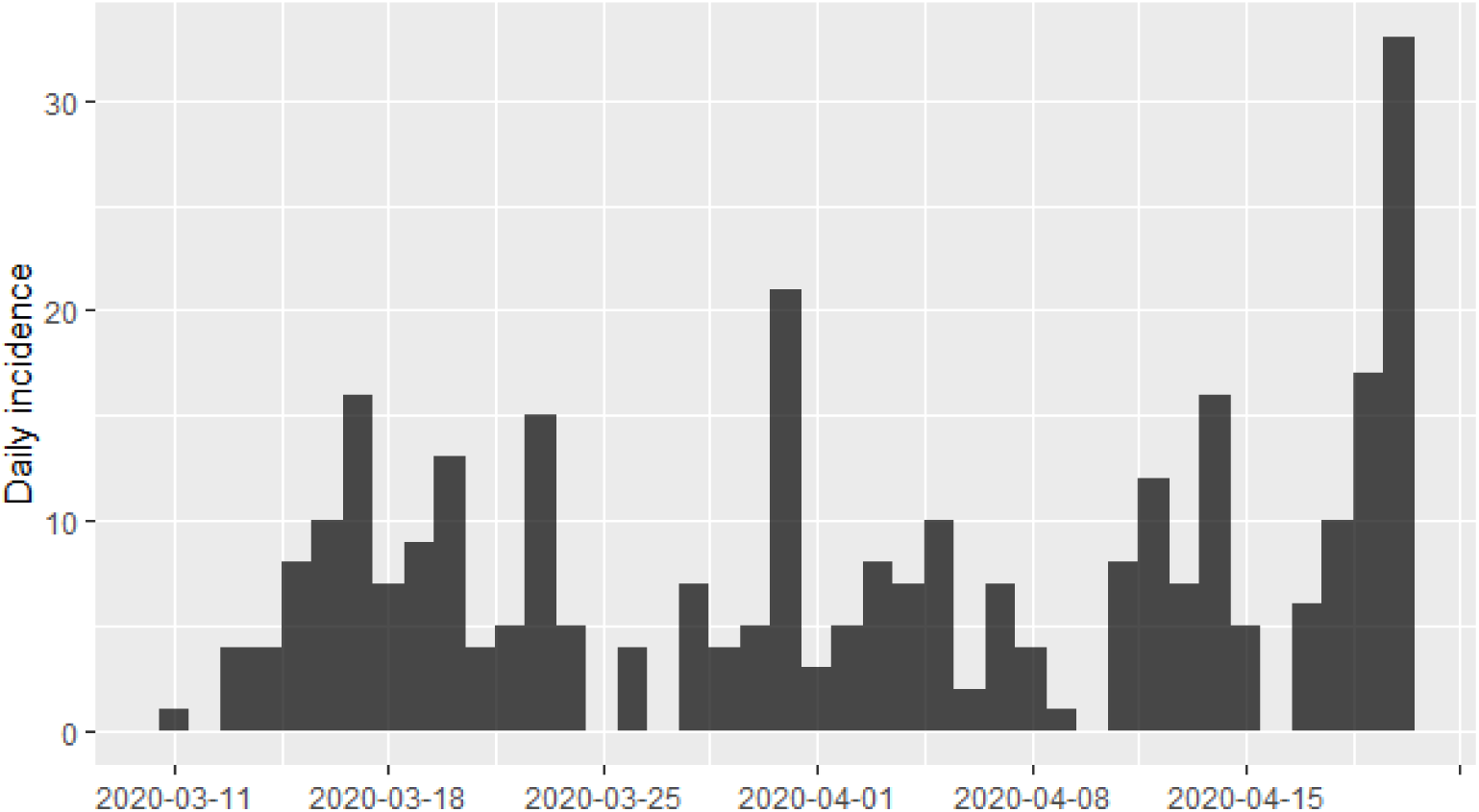
Epidemic Curve of COVID-19 Infections in Sri Lanka

**Figure 14:**
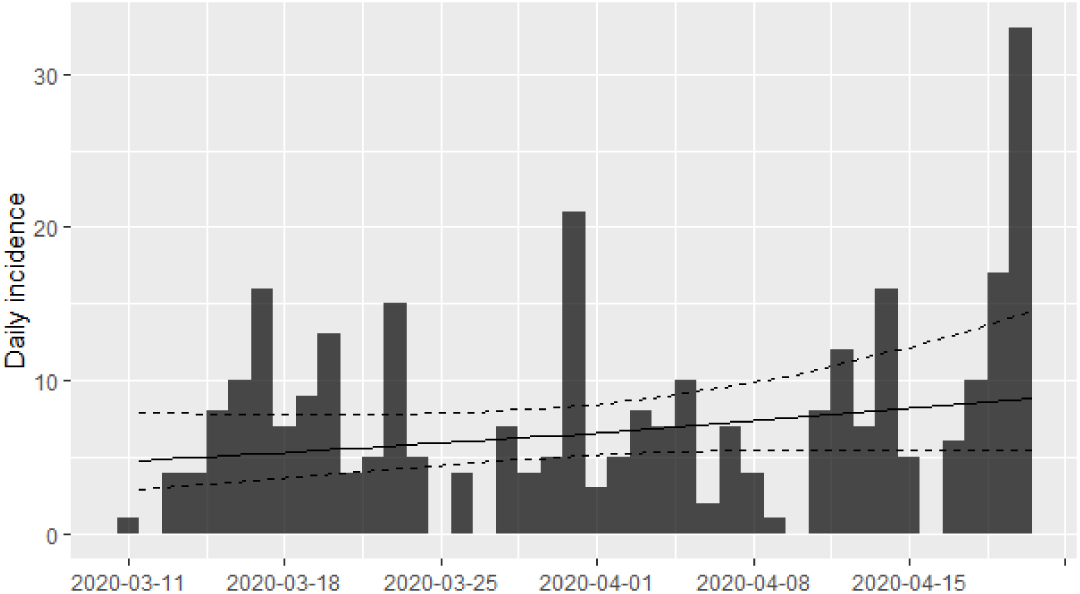
Prediction Model for COVID-19 Infections in Sri Lanka

**Figure 15:**
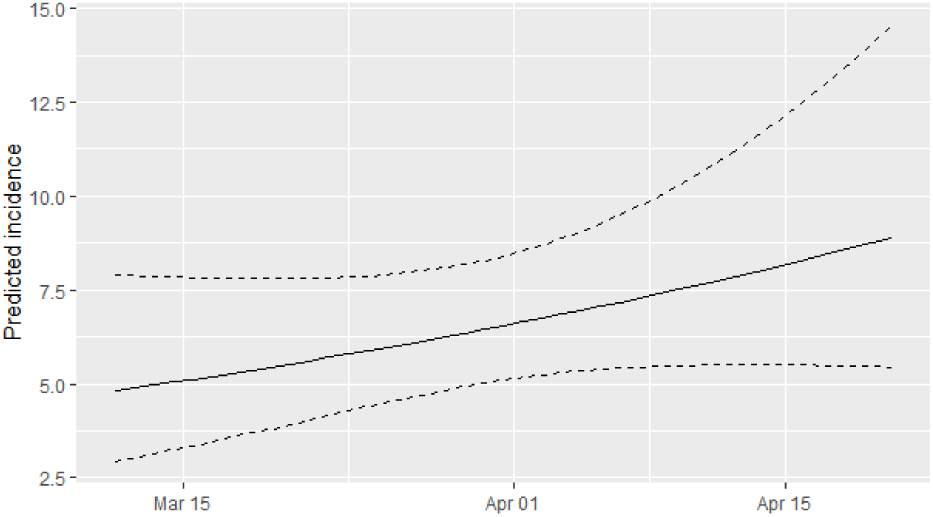
Prediction Models with Intervals

Daily accumulated COVID-19 cases were analyzed using phenomenological models for the countries; Sri Lanka, Italy and Hebei province of China. Particularly, Gompertz, Logistic and Exponential growth curves fitted on cumulative number of infections over the three countries and the best fitted growth curve was identified for each country. The best fitted growth curve had the minimum AIC value. The logistic growth curve was fitted (Figure 16) on the accumulated coronavirus infections reported in Hebei province of China appropriately than the Gompertz and exponential curves by recording minimum AIC measures. The overall growth rate estimated as 4.47. The final epidemic size for the Hebei province of China was estimated as 330 cases. Short-term predictions from the logistic growth curve revealed that the Covid-19 epidemic will reach close to zero in the near future.

**Figure 16:**
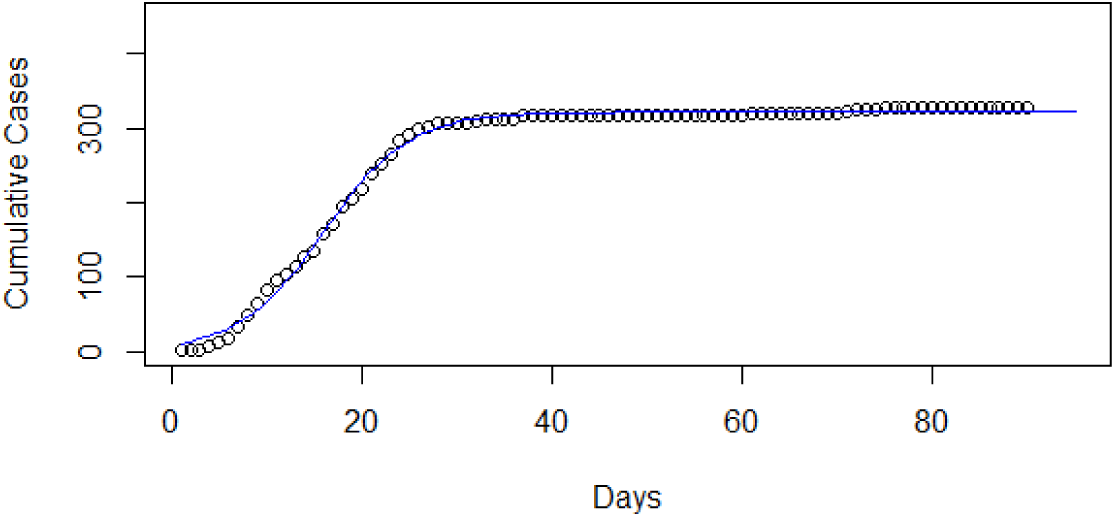
Fitted Logistic Growth Model for COVID-19 infections in Hebei Province, China

The best fitted growth curve for the cummulative number of COVID-19 cases in Italy was a Gompertz growth curve and the fitted curve is shown in Figure 17. When the days increase the number of coronavirus infections decrease at a rate of 0.0692 in Italy. The upper asymptote value was estimated as 2,23,110 cases. Short-term predictions from the Gompertz growth curve revealed that the coronavirus outbreak progressing at a decreasing rate in Italy.

**Figure 17:**
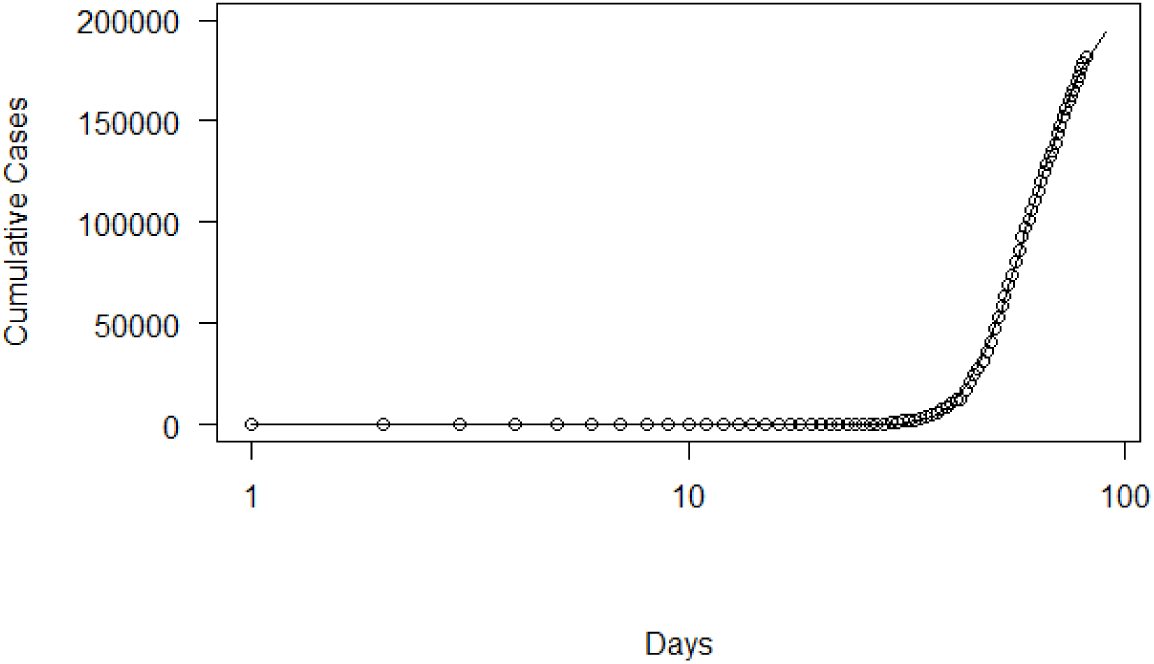
Fitted Gompertz Growth Model for COVID-19 infections in Italy

The most appropriate growth curve for the accumulated number of COVID-19 infections in Sri Lanka was the exponential growth curve which reported minimum value of AIC as 309. The fitted curve depicted in Figure 18. The relative increase of number of infections for a unit increase of days was 0.04. The natural growths at the initial phase follows exponential curves suggested that the epidemic in Sri Lanka is at the initial phase. Short-term predictions from the exponential growth curve revealed that the coronavirus outbreak increasing exponentially in Sri Lanka.1 10 Days

**Figure 18:**
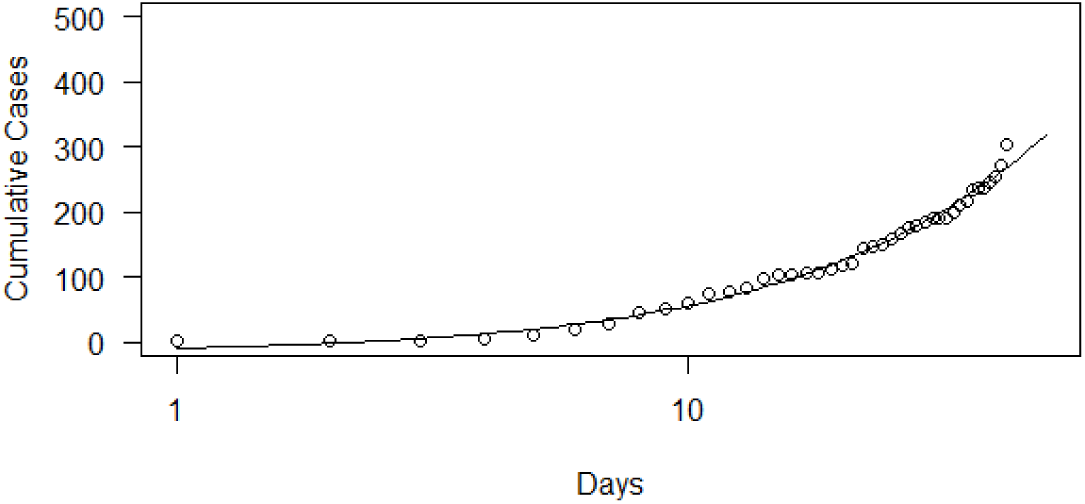
Fitted Exponential Growth Model for COVID-19 infections in Sri Lanka

## 4. Conclusion

This study successfully models the ongoing COVID-19 outbreak present in Hebei province of China, Italy and Sri Lanka using phenomenological modelling and epidemiological curve fitting analysis for the period of date of appearance of cases in each country to the 20^th^ April 2020. The possibility of modelling the outbreak through three growth models; Logistic, Exponential and Gompertz were tested. Findings disclosed that the most appropriate epidemic growth curves for Sri Lanka, Italy and China-Hebei are Exponential, Gompertz and Logistic curves respectively. The epidemiological curves of each country were analyzed to extract useful characteristics of the coronavirus outbreak. Pre-peak and post-peak log -linear models were fitted on epidemic curve to predict the spread of COVID-19 infection in future. The results show that the epidemic seems extinct in Hebei-China whereas further transmissions are possible in Sri Lanka. In Italy, current outbreak transmits in a decreasing rate. Therefore, effective controlling measures need to be continued and enhance with careful supervision in order to keep the epidemic under control in Sri Lanka. In Italy, their controlling mechanisms seems to be on track and should be continued to eradicate the infection.

The analysis of this study was done assuming that all the cases reported accurately. However, there may be under reporting of cases and inadequate screening which leads to a disparity between actual values and the predicted values. Findings of this study will be beneficial for health care professional to allocate necessary resources at the right place at the right time. There is a possibility of applying the methodology used in this study for other countries which has affected with the COVID-19 outbreak to understand/control the epidemic.

## Data Availability Statement

The data used to support the findings of this study are available in HUMANITARIAN DATA EXCHANGE at https://data.humdata.org/dataset/novel-coronavirus-2019-ncov-cases?force_layout=desktop.

